# Ability of Caprini and Padua Risk-Assessment Models to Predict Venous Thromboembolism in a Nationwide Study

**DOI:** 10.1101/2023.03.20.23287506

**Authors:** Hilary Hayssen, Shalini Sahoo, Phuong Nguyen, Minerva Mayorga-Carlin, Tariq Siddiqui, Brian Englum, Julia F Slejko, C. Daniel Mullins, Yelena Yesha, John D Sorkin, Brajesh K Lal

**Author notes:** **Corresponding author and post-publication corresponding author:** Brajesh K Lal MD, Department of Surgery, University of Maryland, 22 South Greene Street, Baltimore, MD 21201.

## Abstract

**Background:** Venous thromboembolism (VTE) is a preventable complication of hospitalization. Risk-stratification is the cornerstone of prevention. The Caprini and Padua are the most commonly used risk-assessment models to quantify VTE risk. Both models perform well in select, high-risk cohorts. While VTE risk-stratification is recommended for all hospital admissions, few studies have evaluated the models in a large, unselected cohort of patients.

**Methods:** We analyzed consecutive first hospital admissions of 1,252,460 unique surgical and non-surgical patients to 1,298 VA facilities nationwide between January 2016 and December 2021. Caprini and Padua scores were generated using the VA’s national data repository. We first assessed the ability of the two RAMs to predict VTE within 90 days of admission. In secondary analyses, we evaluated prediction at 30 and 60 days, in surgical versus non-surgical patients, after excluding patients with upper extremity DVT, in patients hospitalized ≥72 hours, after including all-cause mortality in the composite outcome, and after accounting for prophylaxis in the predictive model. We used area under the receiver-operating characteristic curves (AUC) as the metric of prediction.

**Results:** A total of 330,388 (26.4%) surgical and 922,072 (73.6%) non-surgical consecutively hospitalized patients (total n=1,252,460) were analyzed. Caprini scores ranged from 0-28 (median, interquartile range: 4, 3-6); Padua scores ranged from 0-13 (1, 1-3). The RAMs showed good calibration and higher scores were associated with higher VTE rates. VTE developed in 35,557 patients (2.8%) within 90 days of admission. The ability of both models to predict 90-day VTE was low (AUCs: Caprini 0.56 [95% CI 0.56-0.56], Padua 0.59 [0.58-0.59]). Prediction remained low for surgical (Caprini 0.54 [0.53-0.54], Padua 0.56 [0.56-0.57]) and non-surgical patients (Caprini 0.59 [0.58-0.59], Padua 0.59 [0.59-0.60]). There was no clinically meaningful change in predictive performance in patients admitted for ≥72 hours, after excluding upper extremity DVT from the outcome, after including all-cause mortality in the outcome, or after accounting for ongoing VTE prophylaxis.

**Conclusions:** Caprini and Padua risk-assessment model scores have low ability to predict VTE events in a cohort of unselected consecutive hospitalizations. Improved VTE risk-assessment models must be developed before they can be applied to a general hospital population.

## INTRODUCTION

Venous thromboembolism (VTE), encompassing deep vein thrombosis (DVT) and pulmonary embolism (PE), is a potentially preventable sequala of hospitalization. Over 900,000 VTE events, associated with over 100,000 deaths, and an attendant economic burden of $2-10 billion are reported each year in the United States (US).^1,2^ Summarizing evidence-based guidelines, the US Surgeon General emphasized “the need to screen hospitalized patients for risk of DVT/PE and to provide appropriate prophylaxis to those at risk.”^3^ The Joint Commission on the Accreditation of Healthcare Organizations, the Agency for Healthcare Research and Quality, and the Centers for Disease Control and Prevention have each identified risk-assessment and risk-stratification as the keys to improving VTE prevention in all general hospitalized patients.^4^

At least 23 VTE risk-assessment models (RAMs) have been developed to quantify a patient’s risk for VTE by summing the weights assigned to selected clinical risk-factors.^5^ By mapping ranges of scores into ordered groups, the RAMs attempt to accomplish risk-stratification. The models are designed to be implemented in all hospital admissions, not just high-risk patient groups. The Caprini and Padua RAMs are the most widely used RAMs in hospitals. Risk-factors and weights in both RAMs were selected based on clinical expertise and information from published literature.^6^ The Caprini RAM has been evaluated in high-risk subgroups of surgical patients^7,8^ and the Padua RAM in acutely ill medical patients.^9 10,11^ While both RAMs have been tested primarily in high-risk subsets, guidelines suggest using the Caprini RAM in all hospitalized surgical patients^8,12^ and the Padua RAM in all hospitalized non-surgical patients.^10,11^

Increased awareness about the clinical and cost implications of hospital-associated VTE, and the push to implement risk-stratification by healthcare organizations, has led to an increasing use of the Caprini and Padua RAMs for all hospital admissions. However, neither RAM has been well-studied in a large, unselected cohort of mixed non-surgical and surgical patients.^6,13^

We evaluated the predictive ability of the Caprini and Padua RAMs for 90-day VTE in patients hospitalized at Veterans Affairs (VA) health care facilities nationwide over a 6-year period. In secondary analyses, we assessed predictive performance in subgroups of surgical and non-surgical patients, in patients admitted for ≥72 hours, after including all-cause mortality in the outcome, after excluding upper extremity DVT from the outcome, and after accounting for prophylaxis in the predictive model.

## METHODS

### Study Design and Participants

We performed a retrospective analysis of the first hospital admission of patients to any VA facility nationwide from January 1, 2016, through December 1, 2021. We followed STROBE (Strengthening the Reporting of Observational Studies in Epidemiology) guidelines for cohort studies in writing this report.^14^ Patients with a VTE within 90 days before admission or those admitted with a diagnosis of VTE were excluded. Patients who underwent a surgical procedure (planned or unplanned) during hospitalization, regardless of the admitting service, were defined as surgical patients. Non-surgical patients were those who did not undergo a surgical procedure during hospitalization. The protocol was approved by the Institutional Review Board of the University of Maryland and the Baltimore VA Research and Development Committee.

### Data Source

Data were obtained from the Veterans Affairs Informatics and Computing Infrastructure (VINCI), a comprehensive, patient-level database of the approximately 9 million patients receiving care at 1,298 VA healthcare facilities nationwide.^15,16^ VINCI contains all data entered into the VA’s common electronic medical record from in- and out-patient encounters.

### Outcome and variables

Our main outcome, VTE, was defined as a new International Classification of Diseases (ICD-10) code diagnostic for PE, DVT, or both DVT and PE (**Table S1**). The Caprini RAM computes a composite score based on the sum of weighted scores of 34 risk-factors.^8^ The Padua RAM computes a composite score based on 11 risk-factors.^9^ We computed both scores for each patient using information from their medical record. The risk factors were extracted from various sources: ICD-10 diagnostic and procedural codes, Current Procedural Terminology (CPT) codes, demographic data, clinical and nursing orders, laboratory data, medications, prosthetics consultations, and operating room data. Extracted data included date and time stamps that permitted calculation of length of surgery, duration of bedrest, or date of VTE diagnosis, for example. If there were no admission height or weight measurements, we imputed the body mass index (BMI) using the average of heights and weights obtained within the year before and after admission, after which BMI was available in 96% of patients.

We categorized patients as receiving pharmacologic VTE prophylaxis based on medication orders for unfractionated heparin, low molecular weight heparins, direct-acting oral anticoagulants, or warfarin as well as CPT codes for inferior vena cava filter placement. We determined if patients recieved mechanical prophylaxis based on physician orders for intermittent or sequential compression devices.

### Statistical Methods

We compared demographic and clinical features of our cohort and of the risk-factors included in the Caprini and Padua RAMs for those with and without VTE, using Person’s χ^2^ analysis for categorical variables and Student’s t-test for continuous variables. We used histograms to describe the distribution of RAM scores in patients who did and did not develop VTE, and to describe the relationship between RAM scores and VTE. Logistic regressions in which outcome (yes versus no) was predicted by the Caprini or Padua scores, were used to determine the ability of the two RAMs to predict outcomes of interest and in subsets of interest. Predictive ability was quantified by measuring the area under the receiver-operating characteristic curves (AUC) obtained from the logistic regressions. The AUCs of different models were compared using Delong-Delong tests.^17^ Analyses were performed using R version 4.0.1 (R Core Team, Vienna, Austria) and SAS software version 9.4 (SAS Institute Inc., Cary, North Carolina, USA).

We first evaluated the ability of each RAM to predict VTE in the entire cohort at various time intervals, including 0-30, 31-60, 61-90, and 0-90 days (i.e., four logistic regressions for each RAM). These models adjusted only for the patients’ RAM score. To maximize the number of VTE events included, all subsequent secondary analyses examined outcomes at a single follow-up interval of 0-90 days (our baseline models).

In a series of secondary analyses, we evaluated whether each RAM had better predictive ability (AUC) in various sub-populations of the cohort, or for different outcomes of interest. We first evaluated the prediction of 90-day VTE for each RAM in subgroups of the overall cohort: 1) surgical patients, 2) non-surgical patients, and 3) patients admitted for ≥72 hours, as a surrogate for increased immobility. Next, we evaluated the prediction of 90-day outcome of 4) VTE excluding upper extremity DVTs (because upper extremity DVTs have lower morbidity and mortality) and 5) after including all-cause mortality into the composite outcome (i.e., VTE and/or mortality) since acute PE may remain undetected as a cause of sudden death. Finally, we examined prediction of 90-day VTE by the two RAMs in 6) a subgroup of patients who received VTE prophylaxis (mechanical and/or pharmacologic), 7) a subgroup of patients who did not received VTE prophylaxis, and 8) in the entire cohort after adjusting for any prophylaxis received because prophylaxis may alter the predictive ability of RAMs (prophylaxis is not accounted for in the Caprini or Padua RAMs)

## RESULTS

### Participants

A total of 1,282,014 patients were hospitalized from January 1, 2016, to December 1, 2021. We excluded 21,974 patients admitted with a diagnosis of VTE and 7,580 patients with a VTE within 90 days before admission. Our study population included 1,252,460 patients, of whom 26.4% (n=330,388) were surgical and 73.6% (n= 922,072) were non-surgical patients (**Table 1**). The overall population was older (mean age 65.9 years), predominantly male (93.0%), a substantial minority were non-white (29.8%), and almost 41% received no VTE prophylaxis.

**Table 1:** Clinical characteristics of study cohort

| **Variables** | **All**<br>**N=1,252,460** | **VTE Group**<br>**N=35,557 (2.8%)** | **Non-VTE Group**<br>**N=1,216,903 (97.2%)** | **P-value\*** |
| --- | --- | --- | --- | --- |
| Age, years, mean (SD) | 65.9 (13.8) | 68.1 (12.0) | 65.8 (13.8) | <0.001 |
| Sex, N (%) |  |  |  | <0.001 |
| Male | 1,164,380 (93.0) | 33,846 (95.2) | 1,130,534 (92.9) |  |
| Female | 88,079 (7.0) | 1,711 (4.8) | 86,368 (7.1) |  |
| Race, N (%) |  |  |  | <0.001 |
| White | 891,379 (71.2) | 24,112 (67.8) | 867,267 (71.3) |  |
| Black | 261,905 (20.9) | 8,656 (24.3) | 253,249 (20.8) |  |
| All others | 82,256 (6.6) | 2,250 (6.3) | 82,170 (6.7) |  |
| Body Mass Index, kg/m<sup>2</sup>, mean (SD) | 29.3 (6.8) | 29.2 (7.1) | 29.3 (6.7) | 0.018 |
| Hospital Length of Stay, days, mean (SD) | 4.1 (11.3) | 8.7 (23.5) | 3.9 (10.7) | <0.001 |
| Patient classification, N (%) |  |  |  | <0.001 |
| Surgical | 330,388 (26.4) | 8,064 (22.7) | 322,324 (26.5) |  |
| Non-Surgical | 922,072 (73.6) | 27,493 (77.3) | 894,579 (73.5) |  |
| Prophylaxis Type, N (%) |  |  |  |  |
| Prophylaxis (mechanical and/or pharmacologic) | 740,632 (59.1) | 24,899 (70.0) | 715,733 (58.8) | <0.001 |
| None | 511,828 (40.9) | 10,658 (30.0) | 501,170 (41.2) |  |

\* *P-values computed with Pearson Chi-Square test*
*Abbreviations: SD, standard deviation; VTE venous thromboembolism*

### Outcome data

Within 90 days after admission, 35,557 patients (2.8%) developed a VTE, of which 15,056 (42%) were PEs with or without concurrent DVT, and 20,501 were DVTs alone (58%). The median time (interquartile range, IQR) to VTE occurrence was 11 days (4-35). Although most VTEs developed within the first 30 days of admission, 28% developed 31-90 days later (17% at days 31-60, 11% at days 61-90, **Table 2)**.

**Table 2:**
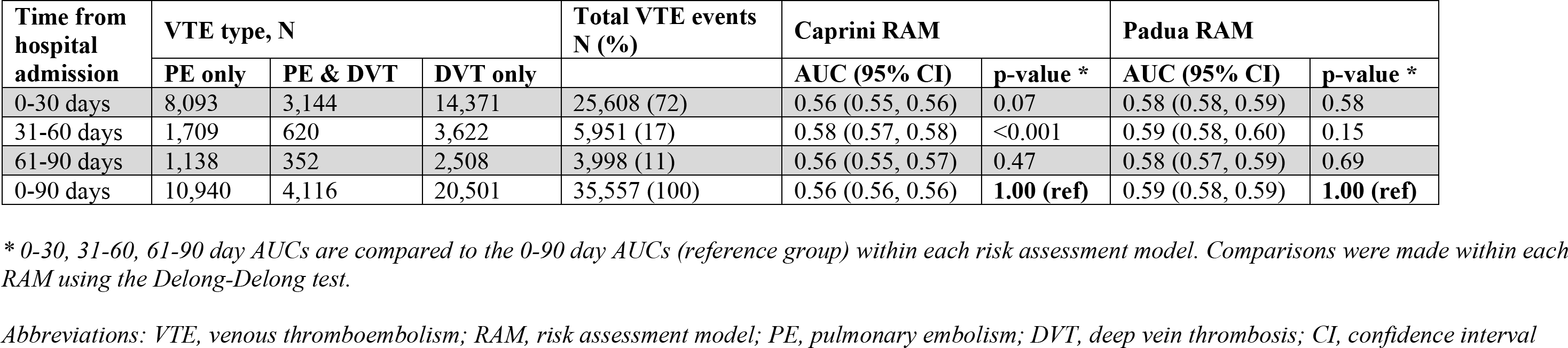
Distribution of VTE events and area under the receiver operating characteristic curves (AUC) for Caprini and Padua scores to predict a VTE after 1,252,460 unique consecutive unselected surgical and non-surgical hospital admissions nationwide.

### Descriptive data (Table 1)

Patients who suffered a VTE (VTE group) were on average older than those who did not suffer a VTE (non-VTE group; 68.1 vs. 65.8 years, p<0.001). Although the difference was not clinically important, the VTE group had a lower BMI than the non-VTE group (29.2 vs. 29.3 kg/m^2^, p=0.018). The fraction who were male was slightly higher in the VTE group than in the non-VTE group (95.2% vs. 92.9%, p<0.001). The fraction who were black was higher in the VTE group than in the non-VTE group (24.3% vs. 20.8% respectively, p<0.001).

Fourteen of the 34 risk-factors in the Caprini RAM were associated with an increased risk of VTE (varicose veins, recent major surgery, swollen legs, heart attack, congestive heart failure, serious infection, lung disease, age 61-74 years, cancer, central venous catheter, age ≥75 years, bedrest ≥72 hours, history of VTE, history of clotting disorder, broken hip, pelvis, or leg, and spinal cord injury resulting in paralysis, **Table 3**). Nine of the 11 risk-factors in the Padua RAM were associated with an increased risk of VTE (infection or rheumatologic disorder, heart attack and/or stroke, heart and/or respiratory failure, age ≥70 years, recent trauma and/or surgery, thrombophilia, bedrest ≥3 days, previous VTE, and active cancer, **Table 3**). Some risk-factors in the Caprini RAM (age 41-60 years, planned minor surgery, inflammatory bowel disease, BMI >25 kg/m^2^, hormone therapy, planned major surgery, and elective arthroplasty) and in the Padua RAM (hormonal treatment, and BMI >30 kg/m^2^) were associated with a reduced risk of VTE.

**Table 3:** Distribution of Caprini ^a^ and Padua ^b^ risk-factors at admission

| **Risk factor** | **Caprini Points** | **Padua Points** | **VTE %**<br>**n=35,557** | **No VTE %**<br>**n=1,216,903** | **p-value<sup>c</sup>** |
| --- | --- | --- | --- | --- | --- |
| Age 41-60 years old | 1 | - | 18.9 | 22.8 | <0.001 |
| Planned minor surgery | 1 | - | 1.3 | 2.7 | <0.001 |
| Varicose veins | 1 | - | 0.4 | 0.2 | <0.001 |
| Recent major surgery | 1 | - | 0.9 | 0.7 | <0.001 |
| Inflammatory bowel disease | 1 | - | 3.3 | 3.7 | <0.001 |
| Swollen legs | 1 | - | 1.2 | 0.6 | <0.001 |
| BMI $\geq 25$ kg/m<sup>2</sup> | 1 | - | 68.4 | 70.7 | <0.001 |
| Heart attack | 1 | - | 4.0 | 2.9 | <0.001 |
| Congestive heart failure | 1 | - | 12.9 | 10.1 | <0.001 |
| Serious infection | 1 | - | 22.3 | 15.5 | <0.001 |
| Lung disease | 1 | - | 16.8 | 16.2 | 0.0015 |
| Bed rest <72 hours | 1 | - | 1.3 | 1.4 | 0.823 |
| Hormone therapy | 1 | 1 | 0.5 | 1.0 | <0.001 |
| Pregnancy | 1 | - | 0.0 | 0.0 | 0.803 |
| History of stillbirth | 1 | - | 0.0 | 0.0 | 0.530 |
| Age 61-74 years old | 2 | - | 52.5 | 48.0 | <0.001 |
| Cancer | 2 | - | 34.7 | 27.7 | <0.001 |
| Planned major surgery | 2 | - | 14.3 | 20.4 | <0.001 |
| Leg cast or mold | 2 | - | 0.03 | 0.04 | 0.764 |
| Central venous catheter | 2 | - | 2.3 | 1.1 | <0.001 |
| Age $\geq 75$ years old | 3 | - | 25.8 | 22.9 | <0.001 |
| Bed rest $\geq 72$ hours | 3 | 3 | 1.4 | 0.6 | <0.001 |
| History of VTE | 3 | 3 | 7.4 | 1.5 | <0.001 |
| Family history of VTE<sup>d</sup> | 3 | - | - | - | - |
| History of clotting disorder | 3 | - | 1.7 | 0.6 | <0.001 |
| Elective arthroplasty | 5 | - | 3.5 | 3.9 | <0.001 |
| Broken hip, pelvis, or leg | 5 | - | 1.6 | 1.1 | <0.001 |
| Serious trauma | 5 | - | 0.0 | 0.0 | <0.001 |
| Spinal cord injury | 5 | - | 0.2 | 0.1 | 0.0010 |
| Stroke | 5 | - | 4.1 | 4.1 | 0.937 |
| BMI $\geq 30$ kg/m<sup>2</sup> | - | 1 | 38.7 | 39.5 | 0.0020 |
| Infection or rheumatologic disorder | - | 1 | 25.7 | 21.1 | <0.001 |
| Heart attack and/or stroke | - | 1 | 6.4 | 5.7 | <0.001 |
| Heart and/or respiratory failure | - | 1 | 16.8 | 11.5 | <0.001 |
| Age $\geq 70$ | - | 1 | 46.6 | 41.2 | <0.001 |
| Recent trauma and/or surgery | - | 2 | 0.9 | 0.7 | <0.001 |
| Thrombophilia | - | 3 | 1.6 | 0.4 | <0.001 |
| Active cancer | - | 3 | 28.4 | 21.1 | <0.001 |

<sup>a</sup> The most recent version of the risk assessment model was used (2013).<sup>8</sup>
<sup>b</sup> The most recent version of the risk assessment model was used (2010).<sup>9</sup>
<sup>c</sup> P-values computed with Pearson Chi-Square test
<sup>d</sup> Unable to extract this information from the medical record; the item was dropped from the computation of the Caprini score
Abbreviations: OR, Odds ratio; VTE, venous thromboembolism; BMI, body mass index; CI, confidence interval

Both Caprini and Padua demonstrated good calibration. The Caprini scores of the cohort ranged from 0-28 (median, IQR: 4, 3-6, **Figure 1**) with an increasing VTE rate as the score increased, a trend that held until a score of 15 (**Figure 2)**. Padua scores ranged from 0-13 (median, IQR: 1, 1-3, **Figure 1**) and an increasing score was also associated with an increasing VTE rate (**Figure 2)**.

**Figure 1.**
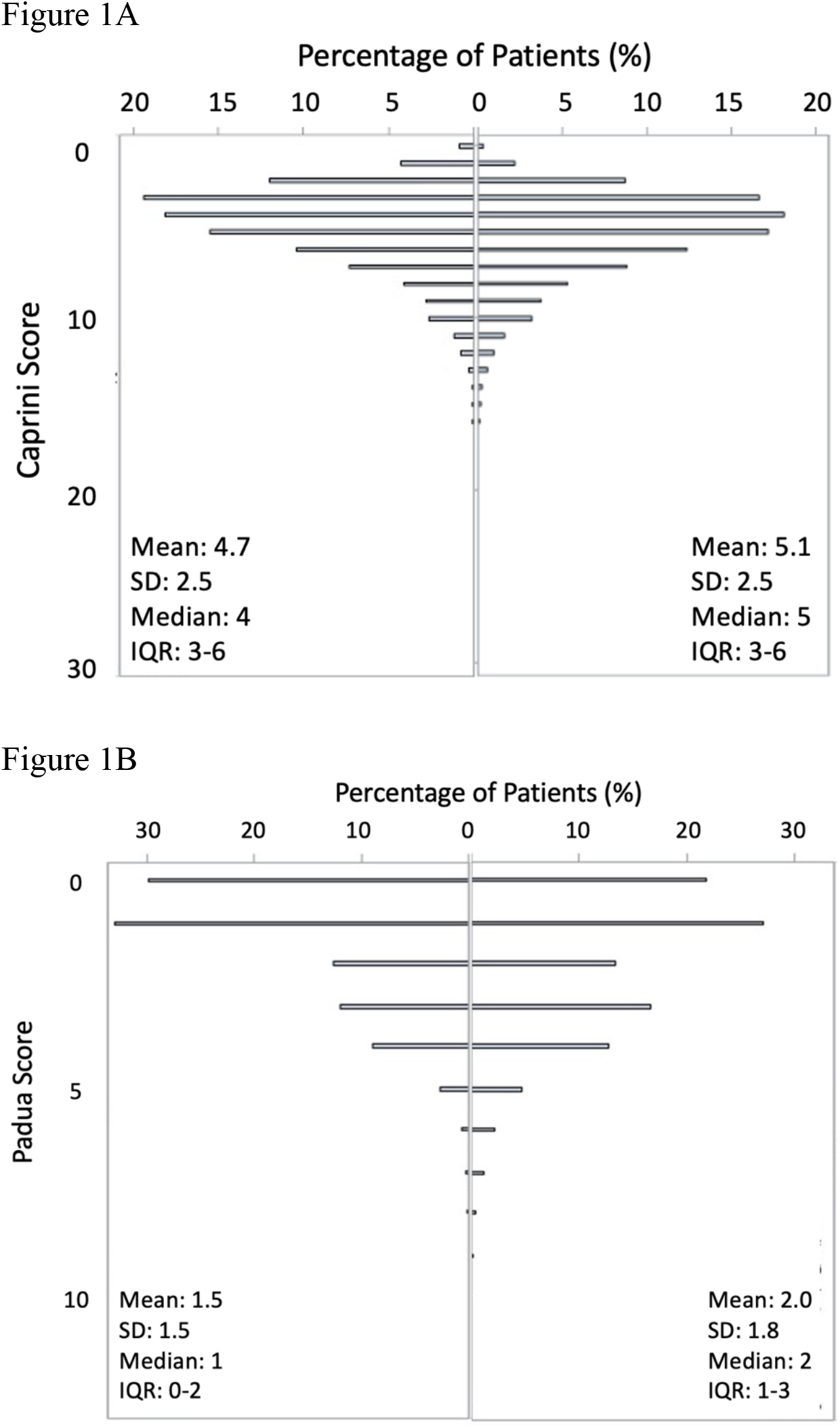
Distribution of (A) Caprini scores and (B) Padua scores at the time of admission from the first admission of 1,252,460 consecutively hospitalized surgical and non-surgical patients. Patient who did not develop a VTE event within 90-days of hospital admission on the **left** and patients that developed a VTE event within 90-days of hospital admission on the **right**. *Abbreviations: VTE, venous thromboembolism; SD, standard deviation; IQR, inter-quartile range*.

**Figure 2.**
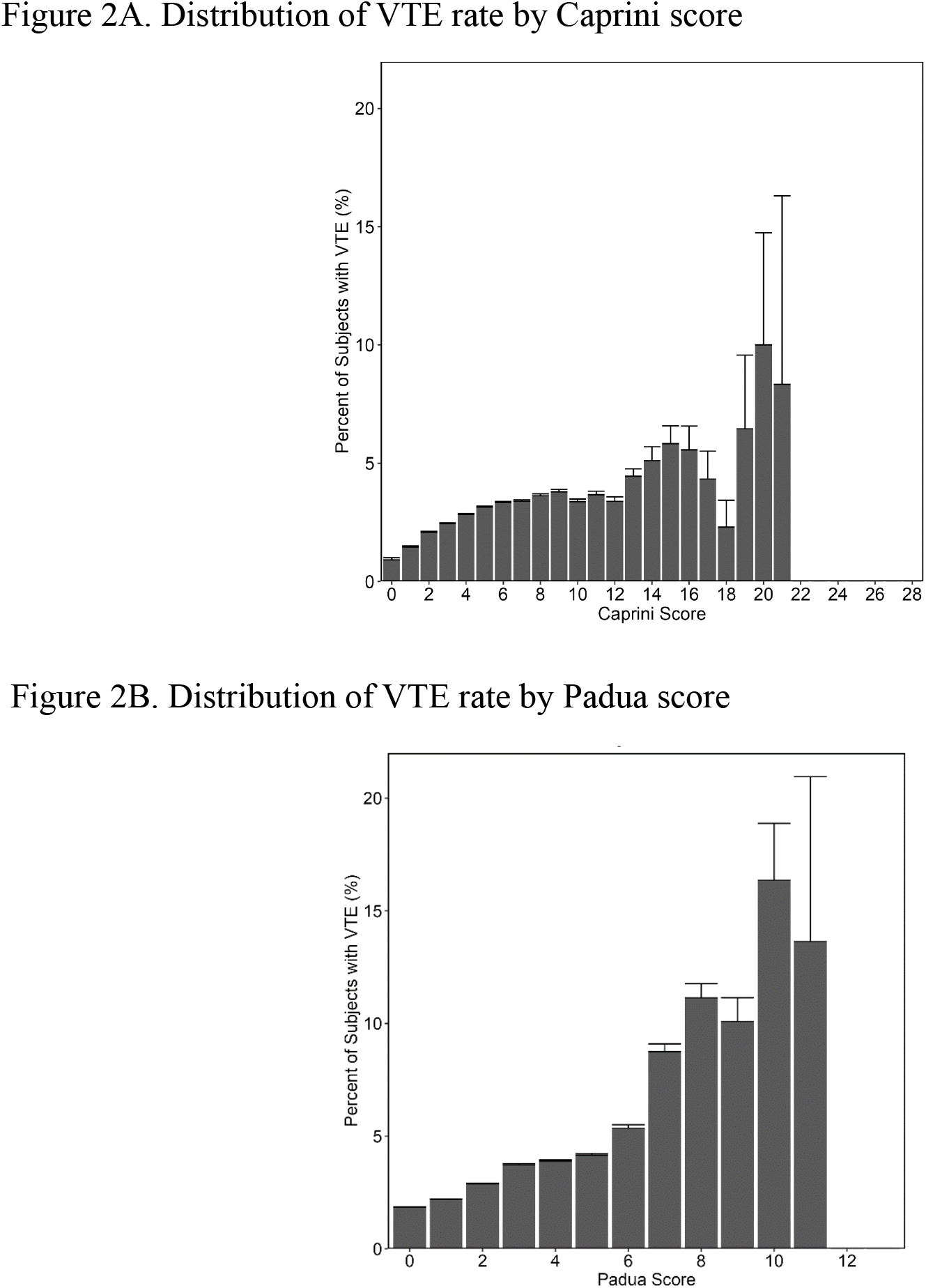
Distribution of VTE rates by risk assessment model score in 1,252,460 consecutively hospitalized surgical and non-surgical patients at the time of their admission. A) Caprini score, B) Padua score. *Abbreviations: VTE, venous thromboembolism; The Error bar represents the upper bar of 95% confidence interval*

### Main results

The AUC for predicting VTE at 0-90 days in the entire cohort by the Caprini RAM was 0.56 (95% CI 0.56-0.56, **Table 2**). The AUCs for 0-30 and 61-90 day outcomes were no different than for 0-90 days, and for 31-60 day outcome was slightly better than for 0-90 days though the difference was not clinically important (absolute difference in AUC, 0.02, **Table 2**).

The AUC for predicting VTE at 0-90 days in the entire cohort by the Padua RAM was 0.59 (95% CI, 0.58-0.59), and was higher than that for the Caprini RAM (p<0.001) though the difference was of limited clinical importance (absolute difference in AUC, 0.03, **Table 2**). The AUCs for 0-30, 31-60 and 61-90 day outcomes were no different from that for 0-90 days (**Table 2**).

### Secondary results (Table 4)

#### Prediction by type of patient examined

In subgroup analyses, the Caprini and Padua RAMs predicted VTE at 0-90 days better in **1)** non-surgical patients (Caprini AUC 0.59 [95% CI 0.58-0.59], Padua AUC 0.59 [0.59, 0.60]) and worse in **2)** surgical patients (Caprini AUC 0.54 [95% CI 0.53-0.54], Padua AUC 0.56 [0.56, 0.57]), compared to VTE prediction in the entire cohort (<0.001 for all four comparisons). Consistent with these findings, prediction in non-surgical patients was better than in surgical patients, though the difference was of limited clinical importance (absolute difference in AUC=0.05, p<0.001, **Figure 3**). The same was observed for the Padua RAM (absolute difference in AUC=0.03, p <0.001, **Figure 3**). Both RAMs performed slightly worse among **3)** patients hospitalized for ≥72 hours versus all patients (Caprini p<0.001, Padua p<0.001).

**Table 4.** Secondary analyses of the Caprini and Padua risk assessment models for venous thromboembolism outcomes within 0-90 days of hospital admission.

| Characteristic Examined | Model # | Analytic approach | Patients analyzed (N) | Events (N) | Caprini RAM |  | Padua RAM |  |
| --- | --- | --- | --- | --- | --- | --- | --- | --- |
|  |  |  |  |  | AUC (95% CI) | P-value<sup>a</sup> | AUC (95% CI) | P-value<sup>a</sup> |
| Predictive ability for VTE in entire cohort (serves as reference for subsequent analyses) | 0 | Full cohort analysis for all bleeding events | 1,252,460 | 35,557 | 0.56 (0.56, 0.56) | 1.00 (ref) | 0.59 (0.58, 0.59) | 1.00 (ref) |
| Predictive ability for VTE by type of patient | 1 | Subgroup analysis of non-surgical patients | 922,072 | 27,493 | 0.59 (0.58, 0.59) | <0.001 | 0.59 (0.59, 0.60) | <0.001 |
|  | 2 | Subgroup analysis of surgical patients | 330,388 | 8,064 | 0.54 (0.53, 0.54) | <0.001 | 0.56 (0.56, 0.57) | <0.001 |
|  | 3 | Subgroup analysis of patients admitted for ≥72 hours | 442,644 | 21,280 | 0.53 (0.52, 0.53) | <0.001 | 0.56 (0.56, 0.57) | <0.001 |
| Predictive ability for different outcomes in entire cohort | 4 | Full cohort analysis for excluding upper extremity DVTs | 1,252,460 | 31,602 | 0.56 (0.56, 0.57) | 0.12 | 0.59 (0.58, 0.59) | 0.57 |
|  | 5 | Full cohort analysis for VTE and/or all-cause mortality | 1,252,460 | 107,684 | 0.58 (0.58, 0.58) | <0.001 | 0.60 (0.60, 0.60) | <0.001 |
| Predictive ability for VTE by prophylaxis status | 6 | Subgroup analysis of patients receiving VTE prophylaxis | 740,632 | 24,899 | 0.55 (0.54, 0.55) | <0.001 | 0.58 (0.57, 0.58) | <0.001 |
|  | 7 | Subgroup analysis of patients not receiving VTE prophylaxis | 511,828 | 10,658 | 0.58 (0.57, 0.58) | <0.001 | 0.59 (0.59, 0.60) | 0.02 |
|  | 8 | Full cohort analysis adjusted for prophylaxis status | 1,252,460 | 35,557 | 0.59 (0.58, 0.59) | <0.001 | 0.61 (0.60, 0.61) | <0.001 |

<sup>a</sup> The AUC of models 1 through 8 were compared to baseline model 0. Comparisons were made within each RAM using the Delong-Delong test.
Abbreviations: RAM, risk assessment model; AUC, area under the receiver operating characteristic curve; CI, confidence interval; DVT, deep vein thrombosis

**Figure 3:**
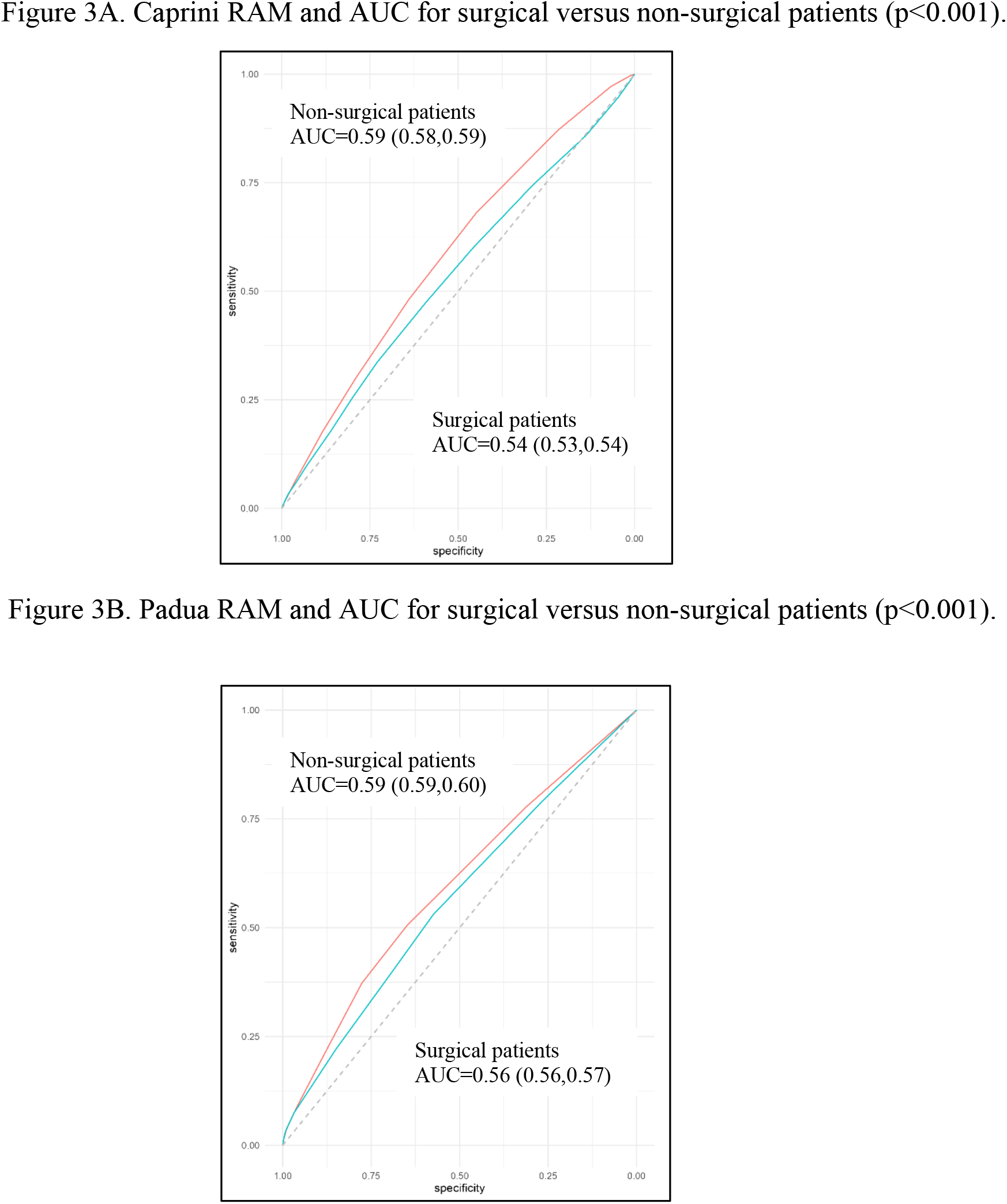
Receiver operating characteristic (ROC) curves demonstrating the ability of the A) Caprini and B) Padua risk assessment models to predict a venous thromboembolism (VTE) event within 0-90 days of hospital admission. Each graph demonstrates the ROC curve for surgical (blue) and non-surgical (red) patients computed separately. *Abbreviations: AUC, area under the curve, the confidence intervals are provided within brackets)*

#### Prediction by type of outcome examined

The AUCs for prediction of **4)** VTE excluding upper extremity DVTs by the Caprini or Padua RAMs were similar to the AUCs for all VTEs (Caprini p=0.12, Padua p=0.57). Both RAMs were slightly better at predicting the **5)** composite outcome of VTE and/or all-cause mortality, versus VTE alone (Caprini p<0.001, Padua p<0.001) although the differences were of limited clinical importance (absolute difference of AUCs, Caprini 0.02, Padua 0.01).

#### Prediction by prophylaxis status

A total of 740,632 (59%) patients received VTE prophylaxis (either mechanical or pharmacologic) and 511,828 (41%) did not; 3.4% of those who received prophylaxis developed a VTE while 2.1% of those who did not receive prophylaxis developed a VTE (p<0.001). 6) In subgroup analyses, for **6)** patients who received VTE prophylaxis, the ability to predict VTE at 0-90 days for both RAMs was worse than for the entire cohort (AUCs: Caprini RAM 0.55 [95% CI 0.54-0.55] and Padua RAM 0.58 [95% CI 0.57-0.58], p<0.001 vs. respective AUCs for prediction in entire cohort). **7)** For patients who did not receive VTE prophylaxis, the ability to predict VTE at 0-90 days for both RAMs was better than for the entire cohort (AUCs: Caprini RAM 0.58 [95% CI 0.57-0.58], p<0.001 and Padua RAM 0.59 [95% CI 0.59-0.60], p=0.02 vs. respective AUCs for prediction in entire cohort). In an additional analysis, **8)** including prophylaxis as an exposure variable in the models for each RAM improved prediction of VTE at 0-90 days compared to our base models (AUCs: Caprini RAM 0.59 [95% CI 0.58-0.59] and Padua RAM 0.61 [95% CI, 0.60-0.61], p<0.001 vs. respective AUCs in unadjusted models, **Table 5**). The differences were of limited clinical importance (absolute differences in AUC, Caprini RAM 0.03; Padua RAM 0.02).

## DISCUSSION

The incidence of VTE within 0-90 days after hospitalization in this nationwide cohort of 1,252,460 unselected consecutive surgical and non-surgical patients was 2.8%. Approximately 28% of VTE events occurred between 31 and 90 days after admission, highlighting the importance of tracking events beyond the traditional focus on 30 days. The ability of the Caprini (AUC 0.56) or Padua (AUC 0.59) risk-assessment models to predict VTE was of limited help in stratifying VTE risk. The performance of both RAMs was better in non-surgical compared to surgical patients, though the improvements were not large enough to be clinically important. The RAMs did not demonstrate clinically important improvements in performance when only patients admitted for ≥72 hours were examined, after excluding upper-extremity DVT from the outcome, after including all-cause mortality into the composite outcome, or after accounting for VTE prophylaxis received. The results do not justify widespread use among general hospital admissions without improvements in the models and further rigorous validation.

Our results confirm that hospital admission confers a risk for VTE. In our study, 2.8% of patients developed a VTE within 0-90 days of hospitalization. Of these, 42% had a PE, an important cause of VTE-related mortality. In addition, with several million patients being hospitalized each year, many patients are at risk for long-term sequelae of VTE such as post-thrombotic syndrome and post-PE syndrome. These iatrogenic complications can be reduced by prophylactic measures. However, mechanical prophylaxis (e.g., compression) may not be tolerated or feasible, and pharmacoprophylaxis may increase the risk for bleeding. These risks must be weighed against the risk for VTE, therefore reliably stratifying VTE risk is a prerequisite to appropriate prophylaxis.

The assessment of predictive performance for the Caprini and Padua RAMs has shown mixed results to date. They are among the most widely used tools to assess VTE risk. The Caprini RAM has been evaluated in over 200 studies worldwide though most have had small sample sizes (<10,000 patients) with few VTE events (<100).^18^ The Padua RAM has been evaluated primarily in medical patients with similarly small cohorts.^19,20^ The best performance (AUCs) for the RAMs were obtained in studies with modest numbers of high-risk patients, often fewer than1,000.^21–23^ The two RAMs have also been compared previously^5,20,23–27^, with the Caprini RAM showing better prediction for mortality,^20^ and a higher AUC to predict VTE (0.77 vs. 0.62 for Padua RAM, p<0.05).^24^ Conversely, an analysis of acutely ill medical patients reported low predictive abilities from both, the Caprini (AUC=0.60) and Padua (AUC=0.64) RAMs.^27^ In our larger, nationwide, mixed surgical and non-surgical cohort, with a large number of VTE events (n=35,557), both RAMs had limited predictive ability for VTE. This may explain why there is limited information on whether broad clinical implementation of the RAMs reduces VTE rates, even though the tools have been available for several years.^28^ A large 43-hospital Michigan Hospital Medicine Safety Consortium instituted systematic VTE risk-assessment in their hospitalized patients using primarily the Caprini RAM. Even though risk-assessment and the use of prophylaxis increased, VTE rates were not reduced.^29^

Most VTE events occurred within 0-30 days, though one-quarter occurred between days 31 and 90 post-admission **(Table 2)**. This could still underestimate the true incidence since PE is often overlooked as a cause of death when it occurs remote from a hospitalization. When we included all-cause mortality into our composite outcome, prediction by both RAMs improved. While the improvement was not large, the increase suggests that some of the deaths occurred in individuals with risk-factors for VTE and could have occurred from a PE. Although we found that hospitalization increased the risk for VTE up to 90 days post-admission, further studies must assess whether risk extends even beyond 90 days, and compare VTE incidence in age-, sex- and risk factor-matched non-hospitalized patients to define the true additional risk for VTE conferred by hospitalization.

Both RAMs showed good calibration with increasing Caprini RAM scores up to a score of 15 being associated with increased VTE rates, at which point the small number of patients and events (and resulting larger confidence intervals) likely contributed to a less linear association **(Figure 2)**. Grant et al. also reported increasing VTE rates up to Caprini scores of 7, after which the rate stayed elevated but without a consistent rise.^30^ Higher Padua scores also demonstrated an increase in VTE rates, consistent with prior reports.^20^

There may be several reasons why the association between RAM scores and VTE events did not translate into better discrimination and prediction of VTE events. Not all risk-factors in the RAMs had a strong relationship to VTE (**Table 3**). In fact, surprisingly, some risk-factors were protective. One explanation may be that both RAMs were empirically derived.^26^ The Caprini RAM was developed using data from only 538 surgical patients, the Padua RAM from only 1,180 non-surgical patients.^7,9^ The prevalence of 13 of the 34 risk-factors in the Caprini RAM were no different between those with versus without VTE, and 5 factors had a higher prevalence in the no-VTE versus VTE group. Future versions of the RAM must reconsider including them in the model.

Obesity is known to have a 2 to 3 fold higher risk for VTE.^31–33^ We found that obesity was not associated with development of a VTE (**Table 3)**. This finding could be related to our study population, older, mostly male, and with many comorbidities, reducing the overall effect of obesity in our cohort. Exogenous hormone use, particularly estrogen, is a known risk factor with a 1.5 to 3 fold higher risk of VTE.^31,34^ In our study, hormone use was more prevalent in the non-VTE group. This is likely a function of the low overall prevalence of hormone use in our population; only 1% of the cohort (n=11,793 patients) had recent (within 30 days) exogenous hormone use, likely due to the small percentage of females (5%).

The Caprini RAM has been tested (and is possibly used) more often in surgical patients, while the Padua RAM has been tested (and possibly used) more often in non-surgical patients. When we assessed for potential differential performance of the RAMs by patient sub-type, we found that both RAMs performed slightly better in non-surgical (AUCs: Caprini RAM 0.59, Padua RAM 0.60) compared to surgical patients **(Table 4)**. The differences were not clinically important. It is possible that some risk-factors potentially relevant to VTE risk in surgical patients are not included in the RAMs (e.g., duration of surgery, type of anesthesia, or organ system being operated on). Since we included all patients that underwent a procedure during hospitalization, regardless of the service they were admitted to, it is unlikely that surgeries were missed in the analysis. The findings argue for an evaluation of a more exhaustive set of risk-factors for inclusion in the RAMs in order to improve performance.

There is no widely used RAM that includes prophylaxis information (that may reduce the risk of VTE), in its calculation of VTE risk.^6^ Studies evaluating the predictive ability of the RAMs have not routinely accounted for ongoing prophylaxis either.^23–25^ We found an improvement in predictive ability of both RAMs when we included information on ongoing prophylaxis into our model (AUCs: Caprini RAM 0.59, Padua RAM 0.61), and when we tested only those patients that did not receive prophylaxis (AUCs: Caprini RAM 0.58, Padua RAM 0.59, **Table 4**). Though the improvements were not enough to make them clinically useful, they suggest the importance of incorporating prophylaxis information into any future model, and into future validation studies. We also found that VTE prophylaxis was associated with a higher risk of VTE. This finding was also noted in a study of 14,660 hospitalized medical patients, where 57% of patients who developed a VTE had received prophylaxis, compared to 46% of those that had not (p<0.001).^27^ This counterintuitive finding may reflect the fact that clinicians, either by calculation of a RAM score, or by clinical intuition, identify patients at high risk for VTE and are more likely to prescribe prophylaxis for these patients. If true, it suggests that the prescribed prophylaxis is not as effective (i.e., appropriate prophylaxis in the appropriate patient) as needed in high risk VTE patients to allow VTE prophylaxis to overcome possible bias by indication.

### Limitations

This is the largest evaluation of VTE RAMs using a nationwide unselected population of general hospital admissions. However, our retrospective study design with reliance on administrative data to define scores of several risk-factors means that Caprini and Padua RAM scores could be under- or over-estimated given inaccurate clinical information. Relying on the VA as the data source decreases the generalizability to female patients and non-VA hospitals. To note though, despite the low fraction of women in our cohort, it is the largest cohort (total number, N=88,079) of women studied for VTE risk-stratification. There is a possibility that clinical data pertaining to event rates from care obtained at non-VA facilities subsequent to hospital discharge may have been missed. However, this would result in underestimating the outcome measure and unlikely to bias our results. Furthermore, the vast majority of Veterans would have returned to VA care and subsequent notes would reflect any events that would have been treated at a non-VA facility. Given the granularity of the VINCI database, we were able to evaluate multiple follow-up periods, different outcomes, and perform several secondary analyses. However, we were not able to include family history in computing the Caprini score as these data were available for only a small fraction of patients. We were missing BMI data on 4% of patients.

While we had data on if mechanical prophylaxis was ordered for patients, we could not confirm patient adherence to the therapy. Possible confounding by indication exists, particularly in the analysis of prediction of VTE in patients receiving and not receiving prophylaxis.

## CONCLUSIONS

The current versions of the Caprini and Padua RAMs have low predictive ability for VTE among general hospital admissions, for both surgical and non-surgical patients. Implementing and ensuring compliance for widespread VTE risk-assessment and risk-stratification using standardized risk-assessment models across large healthcare systems is effort- and resource-intensive. Currently available models must be rigorously assessed and modified before they are ready for universal adoption.

## Data Availability

The United States Department of Veterans Affairs (VA) places legal restrictions on access to veteran's health care data, which includes both identifying data and sensitive patient information. The analytic data sets used for this study are not permitted to leave the VA firewall without a Data Use Agreement. This limitation is consistent with other studies based on VA data. However, VA data are made freely available to researchers behind the VA firewall with an approved VA study protocol. For more information, please visit https://www.virec.research.va.gov.

## Sources of Funding

HH: American Venous Forum (JF2021), T32 (AG000262)

BL: VA Merit Award (RX000995)

JDS: Baltimore VA GRECC (Geriatric Research, Education and Clinical Center), and NIH NIA (P30AG028747)

---

**Table S1:** List of ICD-10 Codes for Diagnosis of Venous Thromboembolism 2

| ICD 10 Code | Description |
| --- | --- |
| I2601 | Septic pulmonary embolism with acute cor pulmonale |
| I2602 | Saddle embolus of pulmonary artery with acute cor pulmonale |
| I2609 | Other pulmonary embolism with acute cor pulmonale |
| I2690 | Septic pulmonary embolism without acute cor pulmonale |
| I2692 | Saddle embolus of pulmonary artery without acute cor pulmonale |
| I2693 | Single subsegmental pulmonary embolism without acute cor pulmonale |
| I2694 | Multiple subsegmental pulmonary emboli without acute cor pulmonale |
| I2699 | Other pulmonary embolism without acute cor pulmonale |
| I81 | Portal vein thrombosis |
| I820 | Budd-Chiari syndrome |
| I82210 | Acute embolism and thrombosis of superior vena cava |
| I82220 | Acute embolism and thrombosis of inferior vena cava |
| I82290 | Acute embolism and thrombosis of other thoracic veins |
| I823 | Embolism and thrombosis of renal vein |
| I82401 | Acute embolism and thrombosis of unspecified deep veins of right lower extremity |
| I82402 | Acute embolism and thrombosis of unspecified deep veins of left lower extremity |
| I82403 | Acute embolism and thrombosis of unspecified deep veins of lower extremity, bilateral |
| I82409 | Acute embolism and thrombosis of unspecified deep veins of unspecified lower extremity |
| I82411 | Acute embolism and thrombosis of right femoral vein |
| I82412 | Acute embolism and thrombosis of left femoral vein |
| I82413 | Acute embolism and thrombosis of femoral vein, bilateral |
| I82419 | Acute embolism and thrombosis of unspecified femoral vein |
| I82421 | Acute embolism and thrombosis of right iliac vein |
| I82422 | Acute embolism and thrombosis of left iliac vein |
| I82423 | Acute embolism and thrombosis of iliac vein, bilateral |
| I82429 | Acute embolism and thrombosis of unspecified iliac vein |
| I82431 | Acute embolism and thrombosis of right popliteal vein |
| I82432 | Acute embolism and thrombosis of left popliteal vein |
| I82433 | Acute embolism and thrombosis of popliteal vein, bilateral |
| I82439 | Acute embolism and thrombosis of unspecified popliteal vein |
| I82441 | Acute embolism and thrombosis of right tibial vein |
| I82442 | Acute embolism and thrombosis of left tibial vein |
| I82443 | Acute embolism and thrombosis of tibial vein, bilateral |
| I82449 | Acute embolism and thrombosis of unspecified tibial vein |
| I82451 | Acute embolism and thrombosis of right peroneal vein |
| I82452 | Acute embolism and thrombosis of left peroneal vein |
| I82453 | Acute embolism and thrombosis of peroneal vein, bilateral |
| I82459 | Acute embolism and thrombosis of unspecified peroneal vein |
| I82461 | Acute embolism and thrombosis of right calf muscular vein |
| I82462 | Acute embolism and thrombosis of left calf muscular vein |
| I82463 | Acute embolism and thrombosis of calf muscular vein, bilateral |
| I82469 | Acute embolism and thrombosis of unspecified calf muscular vein |
| I82491 | Acute embolism and thrombosis of other specified deep vein of right lower extremity |
| I82492 | Acute embolism and thrombosis of other specified deep vein of left lower extremity |
| I82493 | Acute embolism and thrombosis of other specified deep vein of lower extremity, bilateral |
| I82499 | Acute embolism and thrombosis of other specified deep vein of unspecified lower extremity |
| I824Y1 | Acute embolism and thrombosis of unspecified deep veins of right proximal lower extremity |
| I824Y2 | Acute embolism and thrombosis of unspecified deep veins of left proximal lower extremity |
| I824Y3 | Acute embolism and thrombosis of unspecified deep veins of proximal lower extremity, bilateral |
| I824Y9 | Acute embolism and thrombosis of unspecified deep veins of unspecified proximal lower extremity |
| I824Z1 | Acute embolism and thrombosis of unspecified deep veins of right distal lower extremity |
| I824Z2 | Acute embolism and thrombosis of unspecified deep veins of left distal lower extremity |
| I824Z3 | Acute embolism and thrombosis of unspecified deep veins of distal lower extremity, bilateral |
| I824Z9 | Acute embolism and thrombosis of unspecified deep veins of unspecified distal lower extremity |
| I82601 | Acute embolism and thrombosis of unspecified veins of right upper extremity |
| I82602 | Acute embolism and thrombosis of unspecified veins of left upper extremity |
| I82603 | Acute embolism and thrombosis of unspecified veins of upper extremity, bilateral |
| I82609 | Acute embolism and thrombosis of unspecified veins of unspecified upper extremity |
| I82621 | Acute embolism and thrombosis of deep veins of right upper extremity |
| I82622 | Acute embolism and thrombosis of deep veins of left upper extremity |
| I82623 | Acute embolism and thrombosis of deep veins of upper extremity, bilateral |
| I82629 | Acute embolism and thrombosis of deep veins of unspecified upper extremity |
| I82A11 | Acute embolism and thrombosis of right axillary vein |
| I82A12 | Acute embolism and thrombosis of left axillary vein |
| I82A13 | Acute embolism and thrombosis of axillary vein, bilateral |
| I82A19 | Acute embolism and thrombosis of unspecified axillary vein |
| I82B11 | Acute embolism and thrombosis of right subclavian vein |
| I82B12 | Acute embolism and thrombosis of left subclavian vein |
| I82B13 | Acute embolism and thrombosis of subclavian vein, bilateral |
| I82B19 | Acute embolism and thrombosis of unspecified subclavian vein |
| I82C11 | Acute embolism and thrombosis of right internal jugular vein |
| I82C12 | Acute embolism and thrombosis of left internal jugular vein |
| I82C13 | Acute embolism and thrombosis of internal jugular vein, bilateral |
| I82C19 | Acute embolism and thrombosis of unspecified internal jugular vein |
| I82890 | Acute embolism and thrombosis of other specified veins |
| I8290 | Acute embolism and thrombosis of unspecified vein |

